# Mechanical Circulatory Assist Devices Stroke Subtype Classification: A Novel Stroke Classification System in Patients with Ventricular Assist Devices

**DOI:** 10.1101/2025.10.22.25338591

**Authors:** Camila Bonin Pinto, Cameron D. Owens, Ronald Alvarado-Dyer, Andrea Loggini, Jorge Ortiz-Garcia, Mirjam R. Heldner, Shyam Prabhakaran, Faddi G. Saleh Velez

## Abstract

**Background:** Left ventricular assist devices (LVADs) have emerged as the standard of care for bridging-destination therapy in refractory heart failure, however, due to their inherent characteristics, they carry a higher risk of cerebrovascular events. Despite the increased incidence of associated cerebrovascular events in this population, the underlying pathophysiological mechanisms remain poorly understood, hindering effective management and prognosis. We aimed to (1) develop and apply a novel etiological classification system for mechanical circulatory assist device (MCAD)-related strokes, and (2) evaluate the interrater reliability of this classification when applied to cerebrovascular events in LVAD recipients.

**Methods:** The MCAD stroke classification divides LVAD-related strokes in four categories: 1. Management-related (M); 2. Clinically related (C); 3. Acquired (A); 4. Device-related (D). We evaluated the interrater agreement between two independent board-certified vascular neurologists who classified strokes in LVAD patients based on the proposed MCAD classification.

**Results:** We retrospectively reviewed 192 LVAD patients at the University of Chicago Medical Center and identified 32 cerebrovascular events from 2010 to 2020. Among those, 59.4% were ischemic and 40.6% hemorrhagic strokes. The interrater agreement was extremely high (intraclass correlation coefficient = 0.96 ± 0.03). The raters agreed in the primary classification of 32 subjects with the following distribution of mechanisms: 62.6% M, 15.6% A, 15.6% D, 6.2% C. Four subjects fit into more than one category.

**Conclusion:** We propose an MCAD-focused stroke classification that integrates etiology with therapeutic and prognostic decision making. Although evaluated in LVAD patients, this classification can potentially be extrapolated to other mechanical assist devices.

## INTRODUCTION

Stroke is a major cause of disability and a public health concern worldwide. Every year around 795,000 people experience a new or recurrent stroke in the United States with a health care estimated cost of $34 billion.[1] The interest in specific patient subgroups with increased risk for developing strokes, such as patients with left ventricular assist devices (LVADs), has risen in the last two decades. The inherent effects of mechanical circulatory assist devices (MCADs), such as altering hemodynamics and hemostasis, put these patients at an increased risk for various cerebrovascular complications.[2]

An LVAD is a mechanical pump implanted in patients who have reached end-stage heart failure. The device is surgically implanted and is battery-powered. It allows the left ventricle to maintain continuous blood flow to support organ perfusion. While the annual stroke incidence for the general population is less than 1%, the annual incidence of stroke in LVAD patients is estimated to be approximately 8-29% and this variation depends on the LVAD type implanted.[2]

Although there are no large-scale studies addressing the etiology of strokes in this population, it is believed that the cause is multifactorial, with most strokes related to embolism, inappropriate anticoagulation (supra- or sub-therapeutic), and septic emboli that may increase the risk of ischemic or hemorrhagic stroke (HS).[3] Additionally, there is evidence to suggest that the histological composition of the LVAD related emboli is mainly fibrin and denatured protein.[4] However, to date, no data are available regarding the efficacy of thrombolytic therapy (IVT) for this embolus composition. There is also a lack of guidelines and protocols regarding therapeutic goals for blood pressure, anticoagulation resumption, and acute neuro-interventions. In LVAD recipients with large-vessel occlusion (LVO), mechanical thrombectomy (MT) is the primary acute therapy. IVT is generally contraindicated due to anticoagulation. In non-anticoagulated patients meeting criteria, IVT may be administered prior to MT without delaying endovascular treatment,[5-9] leaving most of these patients with a limited therapeutic window for acute reperfusion therapies for ischemic stroke (IS). Similarly, the management of HS is complex, as clinicians must balance anticoagulation reversal, which will increase the risk of LVAD thrombosis.[10]

The underlying pathophysiological and etiologic mechanisms of stroke in this patient population need to be better understood given the heterogenous nature of LVAD related strokes. Accurate differentiation between potential contributory mechanisms can provide critical insight on the most appropriate management strategy. Etiological stroke classifications (e.g., TOAST, ASCOD) have played a pivotal role in understanding and conceptualizing targeted secondary stroke treatments.[11-13] However, none of the current classification systems address patients with LVADs.

In our study, we aimed to analyze and describe our experience in the context of acute strokes in LVAD patients. We analyzed stroke severity, underlying mechanism, adherence to process-based quality measures, acute interventions, and outcomes as to develop a classification system that can guide the clinical management of LVAD patients with cerebrovascular events.

## METHODS

### Study Design

We performed a retrospective chart review from January 2010 to July 2020 at the University of Chicago Medical Center to identify ischemic or HS that occurred at any time point after LVAD implantation. Patients were identified through stroke code activations or inpatient neurology consultations. The diagnosis was confirmed by CT imaging and reviewed by a board-certified vascular neurologist. We excluded patients with transient ischemic attacks, as well as cases where neurological symptoms were attributable to stroke mimics. The study was approved by the University of Chicago institutional review board (IRB20-0778). Informed consent was waived due to the retrospective nature of the chart-review study.

Data collected included patient demographics, baseline clinical status, laboratory results, imaging and diagnostic workup, stroke subtype classification, acute treatments, and outcomes.

### MCAD Stroke Subtype Classification

The MCAD stroke classification categorizes LVAD-related strokes in 4 categories: Management-related (M), Clinically related (C), Acquired (A) and Device-related (D), with subcategories expanded in **Table 1**.

**Table 1.**
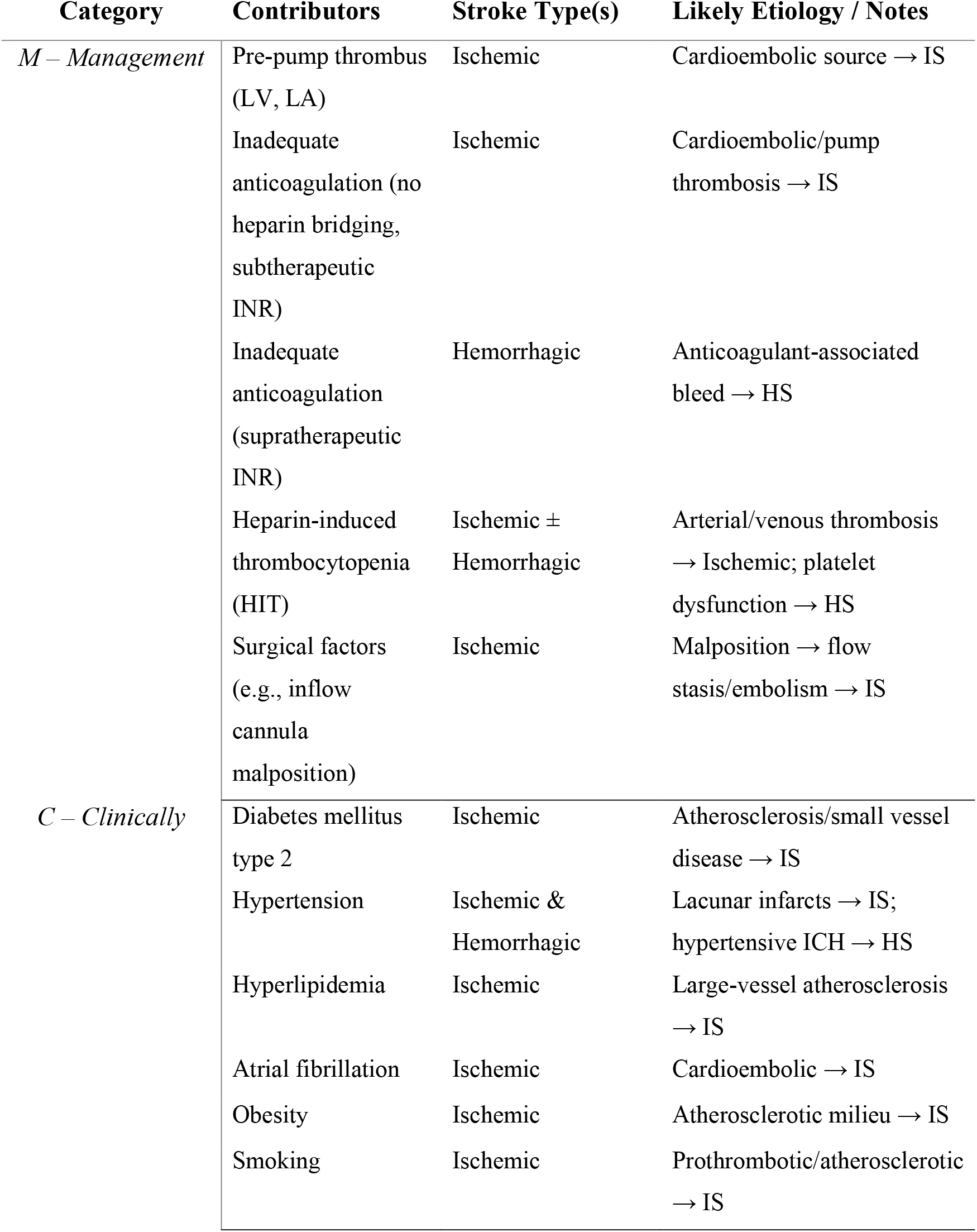
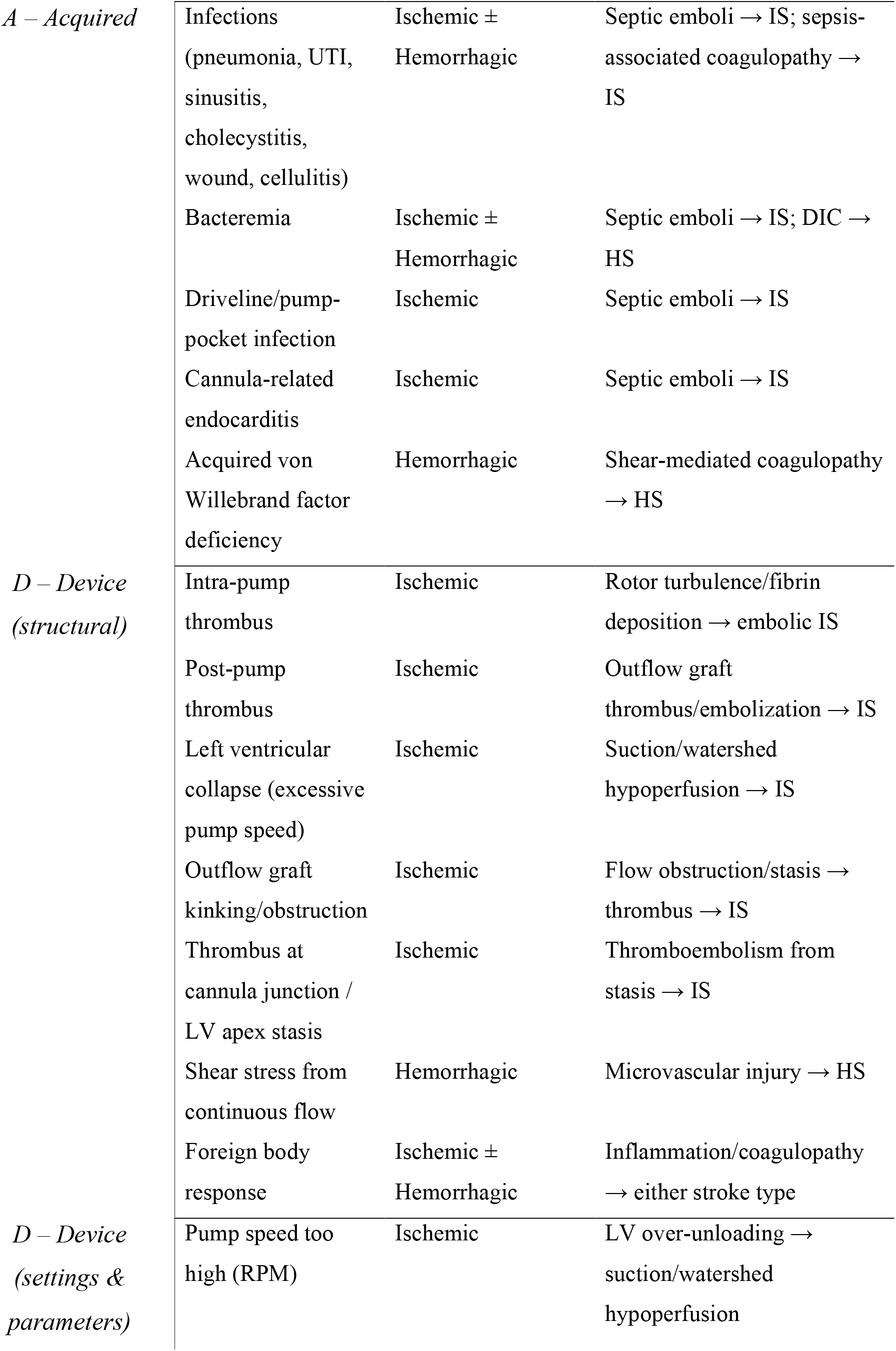
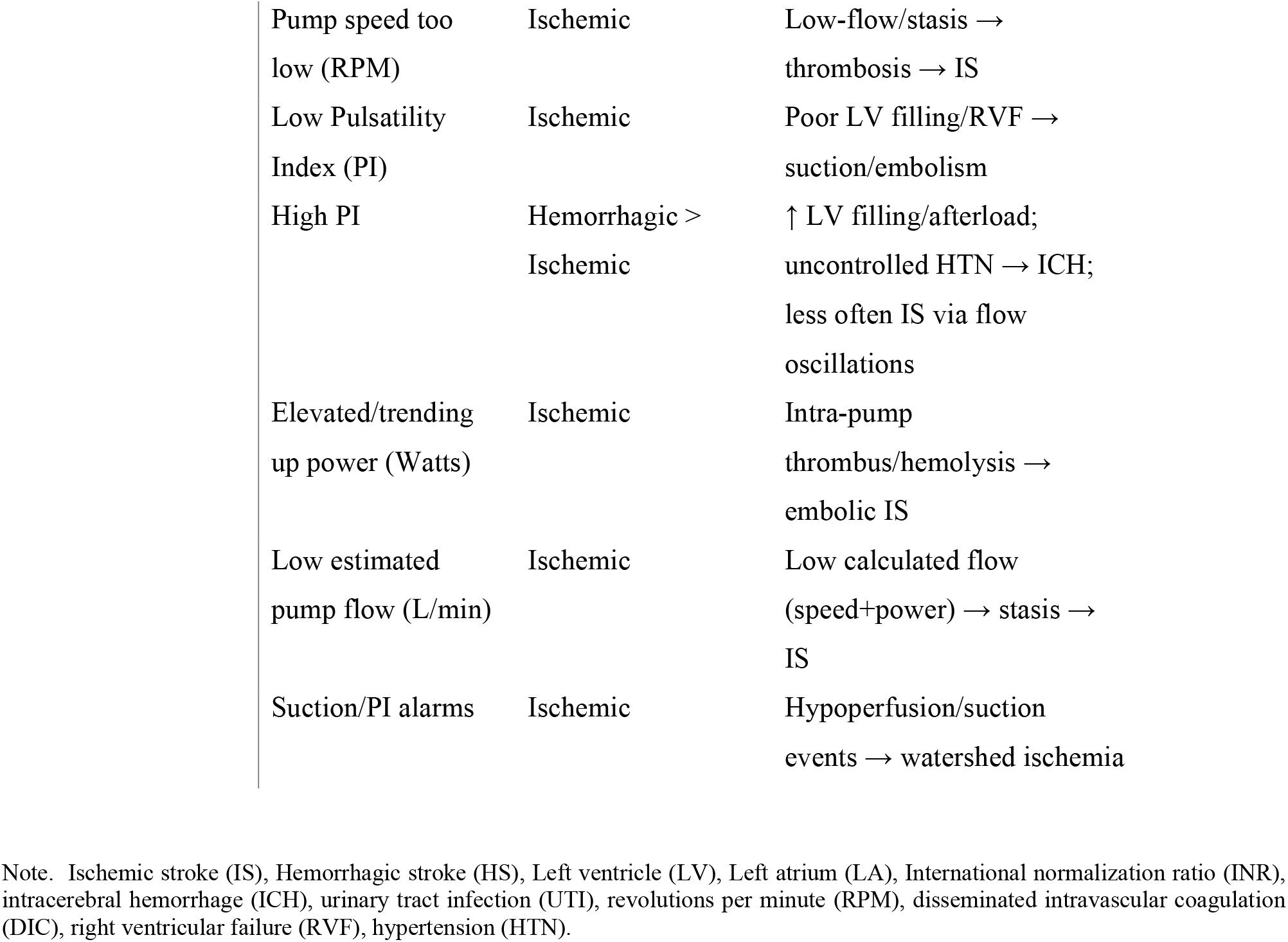
MCAD Classification of LVAD-Associated Stroke, with Stroke Type Attribution and Likely Etiology.

The diagnosis and classification are primarily based on the patient’s clinical characteristics and paraclinical data, including coagulation studies, cardiac imaging, neuroimaging, and LVAD parameter assessments. Given the multifactorial nature of the disease, patients may fit into more than one category, and all contributing factors must be considered essential in guiding therapeutic decisions.

### Testing the MCAD

To test the MCAD classification, two independent board-certified vascular neurologists applied the tool in 32 LVAD patients with either IS or HS at the University of Chicago Medical Center. The rating scale was out of five, with 1 being highly unlikely and 5 highly likely. The two clinicians did not participate in the initial patient encounter but were provided with clinical and paraclinical data of each case. Interrater reliability was used to determine the degree of agreement between the two clinicians to assign each participant to one or more classification groups.

### Statistical Analysis and Data Collection

Charts were reviewed by trained neurology residents who were instructed to gather data already available (e.g., notes, tests, referrals) to identify risk factors and clinical characteristics of the enrolled participants. Characteristics were reported by use of descriptive statistics (e.g., mean for continuous variables, frequency tabulations for categorical variables). Continuous variables were reported as means with standard deviation (SD) when normally distributed and as medians with an interquartile range (IQR) when non-normally distributed. To assess the clinician’s degree of agreement, we calculated inter-rater reliability between the two raters using intraclass correlation coefficient (ICC). The ICC was calculated by dividing the random effect variance (σ2i) by the total variance.[14] The statistical analyses were performed using STATA/SE V. 16.1 (College Station, TX) and IBM SPSS V. 26 (Armonk, NY).

## RESULTS

### Cerebrovascular Events after LVAD: Incidence, Management, and Outcomes

We retrospectively reviewed 192 LVAD subjects identifying 32 (16.6%) cerebrovascular events from 2010 to 2020. Among those, 59.4% were ischemic and 40.6% HS (25% intraparenchymal and 12.5% subarachnoid). Most of the patients were male (65.6%) with a mean age of 56.8 (SD 12.3) and 50% were Black. Ischemic cardiomyopathy was the most common reason for LVAD placement and HeartMate II the most frequently implanted LVAD type. Strokes occurred on average 27.3 months after implantation (25.8 months and 30.9 months to an ischemic and hemorrhagic event respectively).

The majority (n=20) of patients were taking an anticoagulant at the time of the event (62.5%). Among those on anticoagulation, eight combined with aspirin 325 mg, five with aspirin 81 mg and one with aspirin 81 mg and clopidogrel 75 mg a day; leaving 6 subjects with warfarin alone (**Table 2**). As expected, coagulation laboratories revealed supratherapeutic INR more commonly seen in patients that presented with HS, whereas subtherapeutic INR in IS (**Table 3**).

**Table 2.**
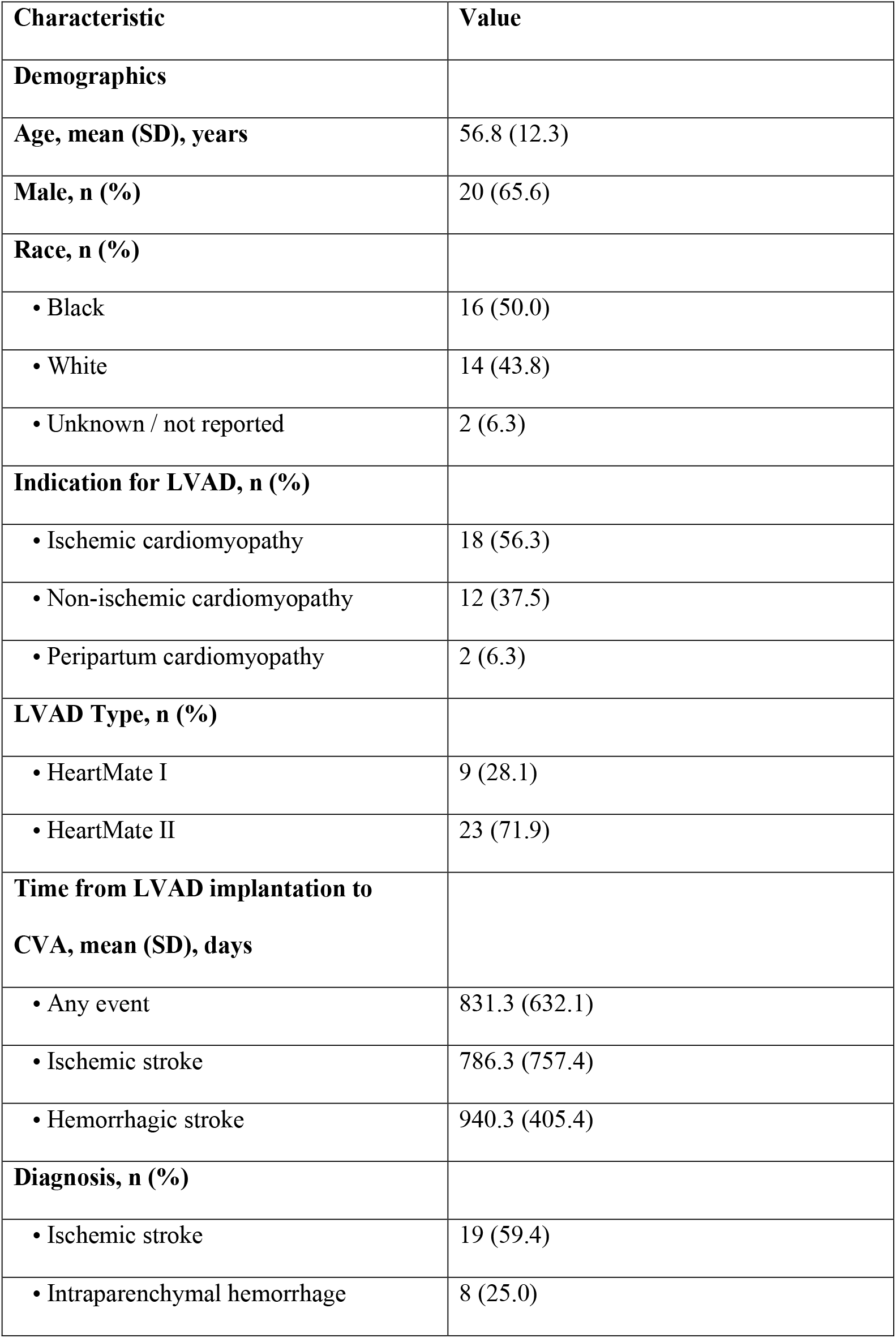
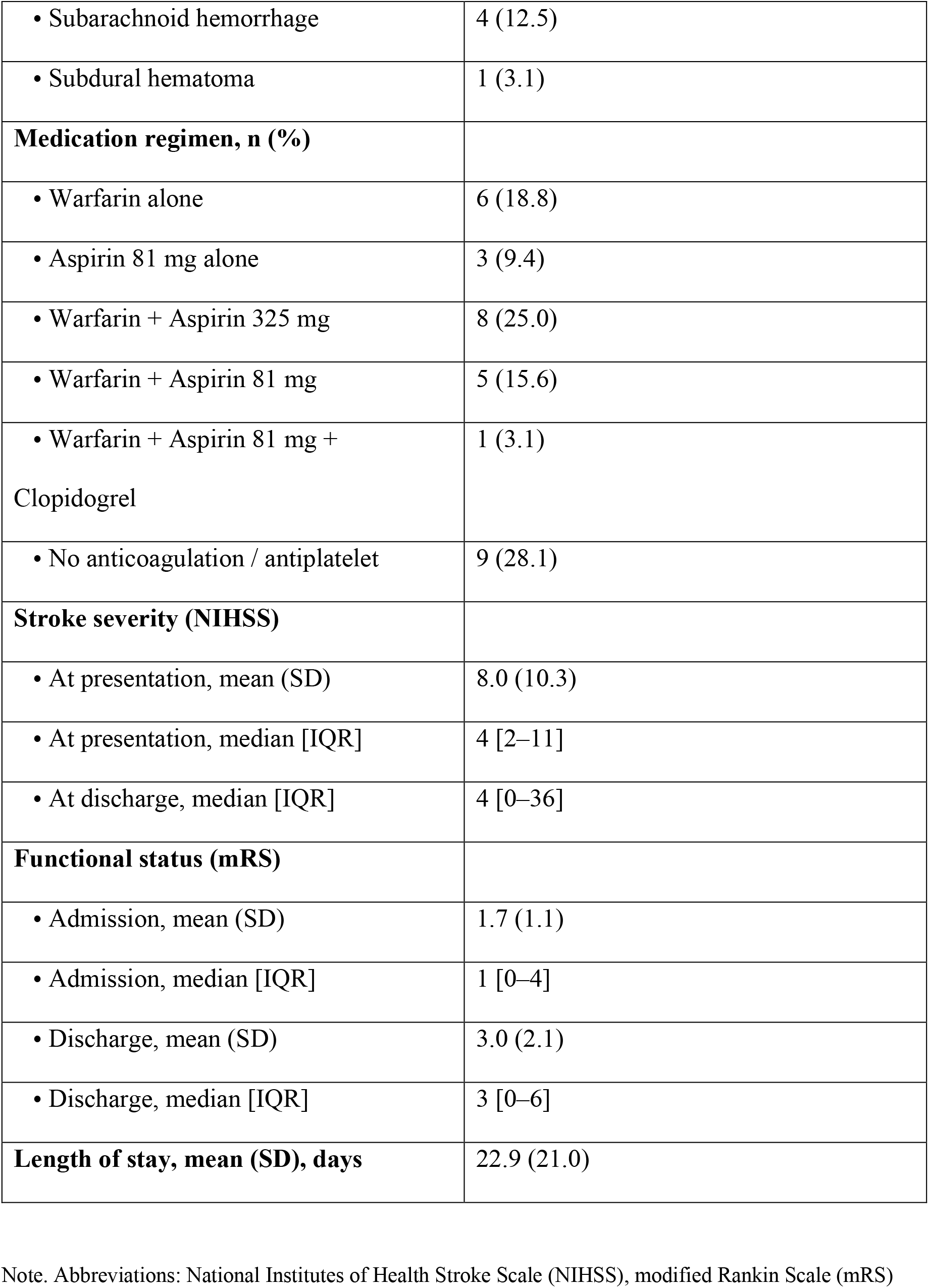
Clinical Profile and Outcomes of LVAD Patients Presenting with Cerebrovascular Events.

**Table 3.**
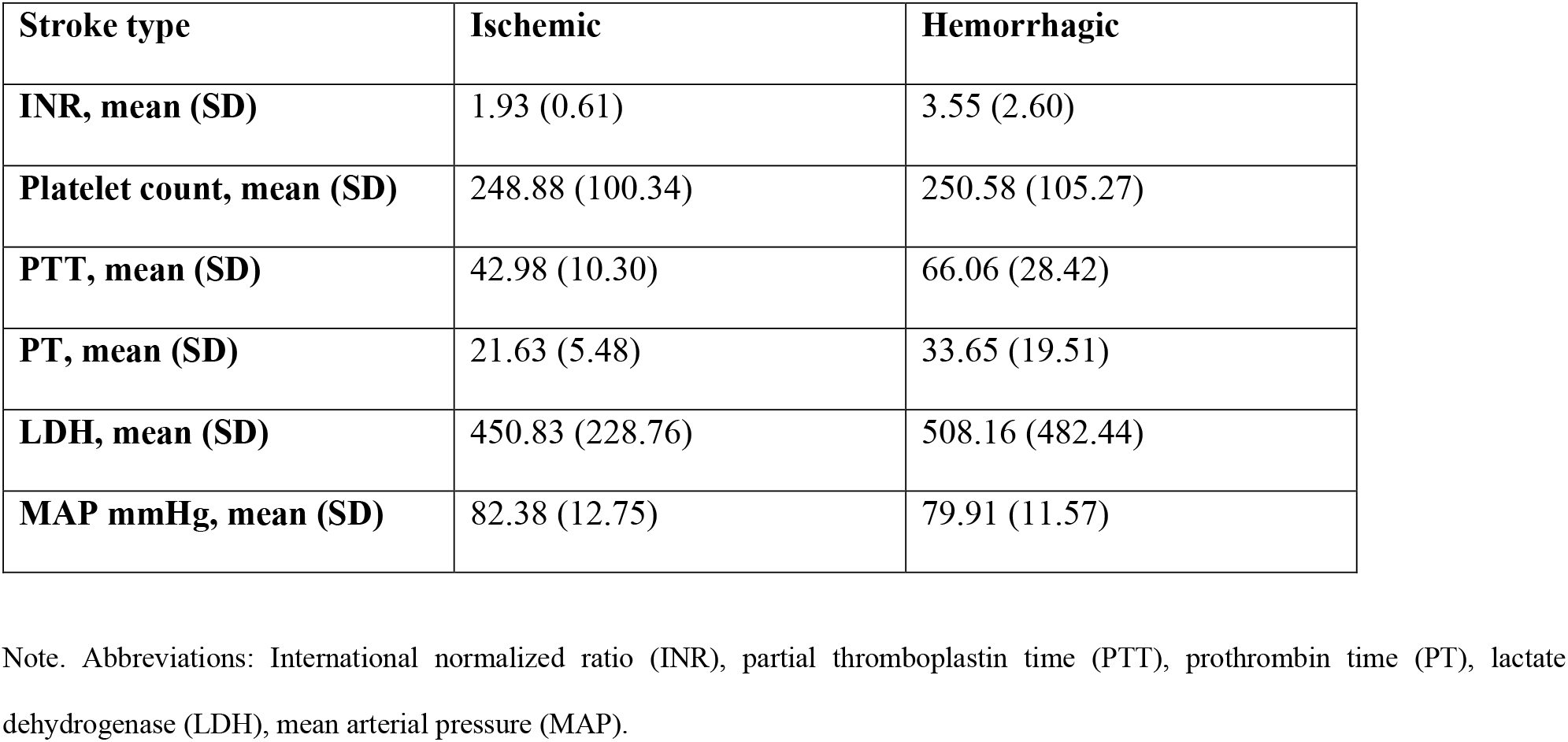
Laboratory Indices and Hemodynamics at Presentation Stratified by Stroke Type.

The most common symptoms were unilateral weakness (37.5%) and altered mental status (e.g., alteration of consciousness, somnolence, confusion, lethargy) (37.5%), followed by aphasia (12.5%), facial droop (12.5%) and headaches (9.37%). The median NIH Stroke Scale at presentation was 4 [IQR 2-11].

None of the 32 subjects received IVT, and only one underwent MT. Medical management was the most used pathway to treat ischemic events with early restart of anticoagulation.

On the contrary, aggressive measurements were required for the case of HS. Multiple means were used to revert anticoagulant states. Among the most common used interventions we found vitamin K (61.5%), prothrombin complex concentrate (53.8%), and fresh frozen plasma (46.2%).

Following IS, there was a median of 1 day to restart antiplatelets [IQR 1 - 1], heparin [IQR 1 - 1.75] and warfarin [IQR 1 - 1.5]. On the other hand, for HS the median for restarting antiplatelets was 7 [IQR 5 - 7] days, and 9 [IQR 7 - 11.75] days for heparin and a more prolonged period waiting to restart warfarin 12 [IQR 12 - 13] days.

After restarting anticoagulation, we found 15.8% of patients had hemorrhagic transformation after IS, however only one was symptomatic, and 7.7% of patients had recurrent bleed following HS. No adverse event was reported after restarting antiplatelet agents.

### MCAD classification

The inter-rater agreement was extremely high (ICC 0.963 ± 0.034) for MCAD stroke classification. The raters agreed in the primary classification of 32 subjects determining a management related etiology in 62.6% cases, and 15.6%, 15.6% and 6.2% for categories A, D and C, respectively. Four subjects fit into more than one category.

For management-related (M) as expected, supratherapeutic INR was the most seen in HS, whereas subtherapeutic INR in IS. Regarding clinically related (C), most of these strokes were lacunar strokes despite having the LVAD. Furthermore, for the acquired (A) group, septicemia was present in 30.8% of the HS and in 5.3% of the IS. Additionally, four HS (46.2%) had a driveline infection and two (10.5%) for the ischemic events. Lastly, for the Device (D) related, we identified 75% (n = 3/5) that underwent a recent visit in which the LVAD parameters were adjusted, and two were found to have a clot during the alert.

### Discharge Disposition, In-Hospital Mortality, and Early Readmissions

Approximately half of patients were discharged home (46.9%) followed by one-sixth to an acute rehabilitation center (15.6%) and one-tenth to a skilled nursing inpatient facility (9.4%). Six patients (18.8%) died during the admission and two (6.3%) were discharged to hospice. A single patient was transferred to an outside hospital per family request. After discharge, the rate of readmission was low, with only two patients readmitted within 30 days and an additional two within the following 90 days.

## DISCUSSION

Strokes in patients with LVADs remain clinically challenging to manage, with limited evidence guiding their presentation, underlying mechanisms, and acute therapy. There are no universally accepted stroke management guidelines specifically tailored for LVAD patients. Decisions regarding acute ischemic therapy are complex. IVT is generally contraindicated due to anticoagulation and perioperative bleeding risk and may be less effective due to the atypical composition of LVAD thrombi (fibrin and denatured protein).[4] MT remains the only viable acute therapy for LVO, although supporting data are limited to small case series.[15] Hemorrhagic events further complicate management, whether as primary intracerebral hemorrhage (ICH) or hemorrhagic transformation, since clinicians must urgently reverse anticoagulation in accordance with ICH guidelines[16] while balancing the high risk of pump thrombosis. To address these gaps and integrate pathophysiology with etiology, prognosis, and therapeutic implications, we developed MCAD, the first stroke classification specifically for patients with MCADs, based on a cohort of LVAD patients **(Figure 1)**.

**Figure 1.**
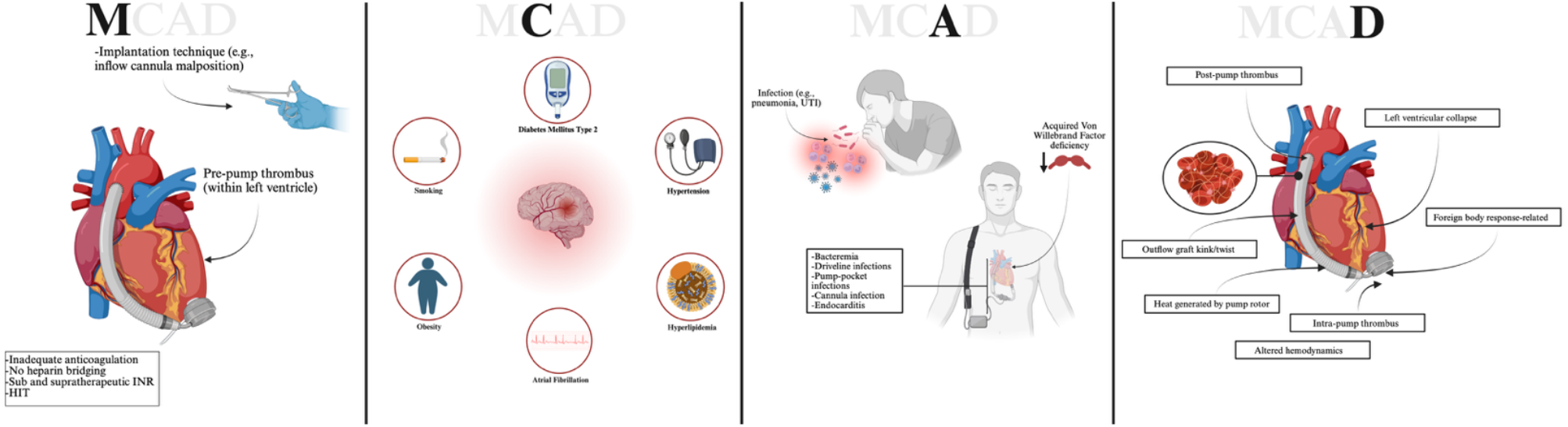
Summary of MCAD classification. Contributors to each category of MCAD (Management (M), Clinically (C), Acquired (A), and Device (D)-related). Created with BioRender.com.

### Management-Related (M)

Antithrombotic management remains central to stroke prevention and treatment in LVADs. Across studies involving HeartMate II, HeartWare, and HeartMate 3, a common strategy is aspirin 81–325 mg daily alongside warfarin targeting INR 2.0–3.0 used for long-term anticoagulation in LVAD patients.[17, 18] However, recent studies favor single anticoagulation, and the HeartMate 3 has demonstrated safety with warfarin alone.[19] Despite these improvements, most patients still receive warfarin plus aspirin, exposing them to additive bleeding risk. Conversely, inadequate anticoagulation increases the risk of thrombus formation, especially in the pre-pump region (left ventricle and left atrium), where impaired contractility and atrial fibrillation (AF) promote blood stasis.[20]

In the perioperative period, unfractionated heparin is commonly used for initial anticoagulation once surgical hemostasis is secured. Heparin infusion is typically started within 12–24 hours after implantation, titrated to achieve partial thromboplastin time goals, and later transitioned to warfarin once the patient is stable. This bridging strategy is critical because delayed anticoagulation initiation markedly increases the risk of early pump thrombosis and IS.[18] Conversely, aggressive or premature heparinization may provoke increased bleeding risk and subsequent HS,[21] underscoring the balance between bleeding and thrombosis. Bridging protocols remain heterogeneous between centers, and no randomized trials have defined optimal timing.

Surgical factors, particularly inflow cannula misalignment, play a notable role in stroke risk among LVAD patients. Mispositioning that creates acute angles toward the septum may disturb flow,[22] creating zones of stasis that predispose to thrombus formation and embolic events. For instance, Schlöglhofer et al. showed that patients with inflow cannula angles <10° had significantly higher rates of IS.[23]

Management-related events underscore the difficulty of balancing bleeding and thrombosis in LVAD patients. In our cohort, IS tended to occur when anticoagulation was subtherapeutic, while HS were more often associated with supratherapeutic states. Moreover, restart of anticoagulation in our cohort (median 1 day for ischemic and 12 days for hemorrhage) contrasts with AHA/ASA guidelines for the general population, which recommend 4–14 days for IS and 4 weeks or greater for ICH.[24] These findings highlight that conventional stroke guidelines may not be directly applicable in LVAD patients, and tailored approaches are required.

### Clinically Related (C)

LVAD recipients typically have extensive vascular risk factors such as hypertension, diabetes mellitus, hyperlipidemia, obesity, AF, chronic kidney disease, and tobacco/alcohol use—all of which amplify stroke risk[1]. Although vascular risk factors were common, clinically related strokes were relatively uncommon in our cohort. Lacunar syndromes were seen less often than expected, likely outweighed by anticoagulation status, acquired and device-related factors. Even so, comorbidities like hypertension, diabetes, and hyperlipidemia still complicate management and warrant ongoing monitoring with labs such as HbA1c, lipid panels, and renal function.

### Acquired (A)

Acquired factors accounted for about one-sixth of classifications in our study. In LVAD patients, infections such as driveline or pump-pocket infections can seed the bloodstream and lead to septic emboli, infarcts, or even mycotic aneurysms that rupture into subarachnoid or intraparenchymal hemorrhage.[17, 25] Acquired von Willebrand disease, driven by shear-mediated cleavage of high-molecular-weight multimers, also increases bleeding risk and is well recognized in continuous-flow devices.[26] Together, these acquired contributors highlight the importance of attention to infection and bleeding syndromes in this population.

### Device-Related (D)

Device-related contributors reflect both structural factors and real-time pump parameters. Thrombus can form inside the pump itself, where shear stress and turbulence within the rotor housing promote fibrin deposition, power spikes, hemolysis, and risk of embolic stroke.[7, 27] Post-pump complications, such as outflow graft kinking, obstruction, or thrombus deposition, likewise predispose to systemic embolization or hypoperfusion.[28] Other structural risks include thrombus at the cannula junction and blood stagnation at the left ventricular apex, both of which may lead to ischemic emboli.[29]

Pump console parameters provide important early warning signs. Elevated power usually signals intra-pump thrombosis,[30] while low flow may reflect hypovolemia, right ventricular failure, or graft obstruction.[31] High pump speed can over-unload the LV, producing suction events and watershed ischemia, whereas a low pulsatility index (PI) indicates poor LV filling and ischemic risk.[32, 33] Conversely, high PI often reflects increased afterload (e.g., hypertension), indirectly raising HS risk.[34] Because afterload critically modulates LVAD hydraulics, aggressive blood pressure control has emerged as one of the few modifiable device-related stroke risk factors. In the ENDURANCE Supplemental trial, strict blood pressure management significantly reduced stroke rates in HVAD recipients.[35]

In our cohort, device-related events accounted for a meaningful subset of cases, frequently occurring after recent LVAD parameter adjustments or in the setting of confirmed device thrombus. These mechanisms underscore the importance of routine pump log review, hemodynamic optimization, and vigilant blood pressure management after neurological events in LVAD patients.

### Interrater Agreement

We found high interrater agreement in applying the MCAD classification, suggesting the system is both replicable and clinically intuitive. Importantly, it guides clinicians to consider a comprehensive checklist of anticoagulation, comorbidities, infection, and device parameters when evaluating stroke etiology in LVAD patients. This structured approach not only improves diagnostic accuracy but also facilitates a rational plan for antithrombotic resumption, infection management, and device troubleshooting.

### Limitations and Future Directions

Our study has several limitations. Its retrospective nature limited assessments to what was documented in charts. The single-center setting and modest sample size restrict generalizability. However, strengths include the systematic review of laboratory values, LVAD parameter logs, vital signs, and imaging by two independent assessors, which enhances confidence in our findings. Future directions include validation of the MCAD classification in larger, multicenter cohorts, as well as prospective studies to refine timing of antithrombotic resumption, integrate device parameter monitoring, and evaluate whether applying this framework can improve stroke outcomes in LVAD patients.

## CONCLUSION

Using the MCAD stroke classification, two independent reviewers categorized all 32 cerebrovascular events, demonstrating strong interrater agreement. The most frequent etiologies were management-related anticoagulation issues, followed by device-related factors, acquired complications of long-term support, and clinically-related factors. This framework provides a structured approach to evaluating strokes in LVAD patients, standardizes diagnostic workup, and guides both acute and secondary prevention strategies, ultimately enabling more individualized patient care.

## Ethical Approval and Informed Consent Statements

Approved by the University of Chicago Institutional Review Board (IRB20-0778); in accordance with the Declaration of Helsinki; requirement for informed consent was waived.

## Data Availability Statement

De-identified data and the analysis code are available from the corresponding author on reasonable request

